# Methylation-associated Gene Expression (MaGE): a non-invasive, in vivo measure of cellular senescence

**DOI:** 10.64898/2026.09.29.26364335

**Authors:** Daniel J. Simpson, Samuel J. Crofts, Christos Mavrommatis, Thomas A. White, Yossawat Suwanlikit, Kirstyn Loyva, Walter A. Rocca, Kejal Kantarci, Michelle M. Mielke, Anne Richmond, Riccardo E. Marioni, Kristina Kirschner, Nathan K. Lebrasseur, Tamir Chandra

## Abstract

Cellular senescence is a central hallmark of aging, yet its measurement in humans remains invasive, low-throughput and tissue-specific, precluding population-scale study. We developed Methylation-associated Gene Expression (MaGE) predictors: whole-blood DNA methylation proxies of p14^ARF^, p16^INK4A^ and p21^CIP1^ expression, plus a composite score, providing the first scalable measure of senescence-marker expression. Deployed across Generation Scotland (n=18,859), MaGE yielded 45 Bonferroni-significant associations spanning 18 incident diseases and all-cause mortality. The two strongest associations recapitulated the established tissue specificity of p21^CIP1^ and p16^INK4A^ activation, with MaGE-p21 associating with incident alcoholic liver disease and MaGE-p16 with pulmonary fibrosis. In a separate study, MaGE tracked disease severity and treatment response in Crohn’s disease. MaGE is a scalable, interpretable biomarker of senescence that enables its study at population scale and offers a route to patient stratification in senolytic trials.

## Introduction

Since the discovery of cellular senescence as a state of stable growth arrest ^1^, approaches to identify and characterize senescent cells have evolved from individual markers, such as *p16^INK4A^* (*CDKN2A*) ^2^ and senescence-associated β-galactosidase ^3–7^, to multiomic strategies that capture the multidimensional nature of the senescent phenotype ^8–10^. These approaches have enabled increasingly refined definitions of senescent cell states (“senotypes”) in culture and across tissues in preclinical models of aging and disease ^8–10^. Yet, despite these advances, our ability to quantify senescent cell burden in humans remains limited, which precludes the investigation of its direct contribution to aging and age-related diseases, which (despite support from senescent-cell ablation studies in mice ^11,12^) remains unresolved in humans. Tissue-based approaches are invasive, difficult to scale and poorly suited to repeated sampling ^3^, creating a critical need for blood-based biomarkers that can quantify senescent cell burden and enable its longitudinal assessment in living individuals ^13,14^.

Among candidate blood-based indicators of senescence burden, *p16^INK4A^* expression in CD3+ T cells has substantial biological and clinical support. CD3+ T-cell *p16^INK4A^* expression increases with chronological age and is associated with exposures and interventions linked to biological aging and cellular stress, including smoking, physical activity, caloric restriction, inflammation, chemotherapy, hematopoietic stem cell transplantation and HIV infection ^2,13–22^. Despite this body of evidence, its measurement requires fresh blood, cell isolation, and technically demanding processing. These barriers limit its utility for routine clinical practice, large-scale cohort studies, and retrospective analyses of archived biospecimens.

In contrast, DNA methylation (DNAm), can be easily measured at scale and provides a versatile platform for blood-based biomarker development. DNAm-derived measures are well established as indicators of biological aging, mortality and disease outcomes ^23–31^, and can also serve as surrogates for circulating molecules, including inflammatory proteins ^32–41^. A categorical strength of DNAm-proxy technologies is that a single DNAm assay can support multiple existing and emerging metrics and can be retrospectively applied to existing large-scale population cohorts, such as Generation Scotland (GS) ^35–40,42^. This creates an opportunity to develop DNAm-based indicators of senescence burden that can be readily deployed across existing and prospective clinical and population-based cohorts.

Here, we developed Methylation-associated Gene Expression (MaGE) predictors to provide scalable blood-based indicators of cellular senescence. Using whole-blood DNAm, we trained predictors of transcript abundance for three key senescence-associated genes, *CDKN2A* (p16^INK4A^ and p14^ARF^) and *CDKN1A* (p21^CIP1^) as measured in the established CD3+ T-cell p16^INK4A^ expression assay ^2^. We applied these predictors, individually and as a composite measure, to 18,859 participants in GS ^42^ and examined their associations with incident disease and mortality, their performance in independent disease cohorts, and their genetic determinants. Together, these analyses establish the potential of MaGE to translate molecular indicators of cellular senescence into scalable measures that can be applied across large human populations and existing DNAm datasets.

## Results

### Development of MaGE to predict in vivo senescence-associated transcript abundance

An overview of the workflow is given in Figure 1. First, we analyzed DNAm patterns in whole blood DNA and gene expression in isolated CD3+ T-cells obtained from 454 women aged 55-82 years in the Mayo Clinic Cohort Study of Oophorectomy and Aging-2 (MOA-2) ^43^ cohort (clinical characteristics provided in Supplemental Table 1). We then trained DNAm predictors of p16^INK4a^, p14^ARF^, and p21^CIP1^ (Fig.1A). Candidate CpGs were selected by mutual information with expression of each gene (Fig.1B), and elastic net regression was used to generate the three individual MaGE predictors, alongside a composite MaGE score (Fig.1C). We then applied these predictors at scale in a large healthy population cohort (Generation Scotland, n=18,859), assessing their association with 174 incident disease outcomes and benchmarking them against established epigenetic clocks and routinely collected clinical measures, before performing a GWAS of all four MaGE predictors. Finally, we evaluated MaGE predictors in independent disease-specific DNAm datasets of COVID-19 and Crohn’s disease (Fig.1D), considering their associations with disease severity and resolution.

**Figure 1.**
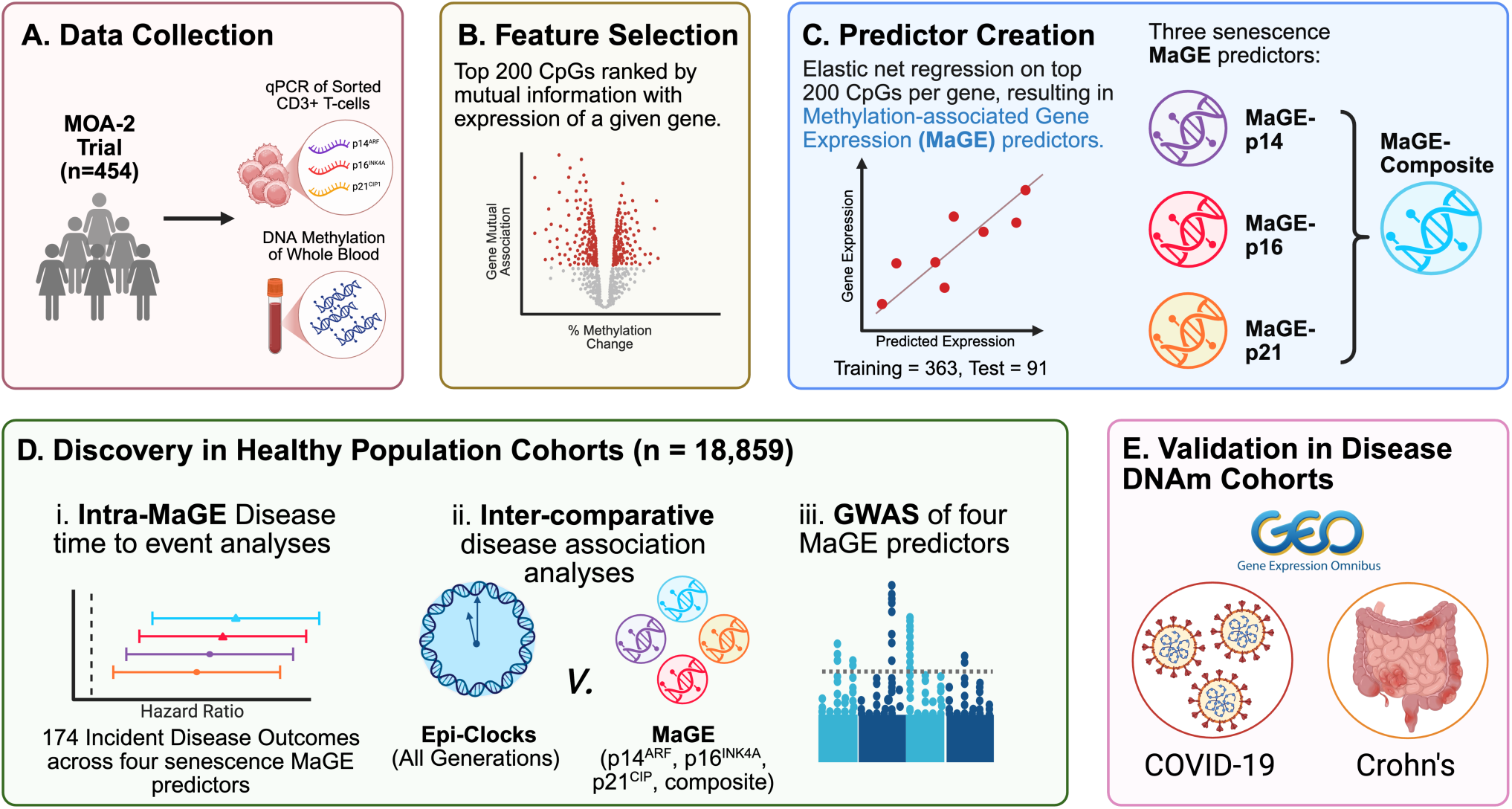
Study Overview. **A.** Paired DNAm and qPCR expression data were obtained from the Mayo Clinic Cohort Study of Oophorectomy and Aging-2 (MOA-2) cohort (n=454)^43^. **B.** Feature Selection identified 200 CpGs via MI. **C.** We computed elastic net regressions on the 200 selected CpGs, creating 3 distinct MaGE proxies, as well as their composite **D.** The four predictors were then assessed within Generation Scotland as risk predictors of 174 incident diseases and all-cause mortality, benchmarked against standard clinical markers and established epigenetic clocks. GWAS of MaGE predictors identified hits at senescence-related genes potentially regulating p14^ARF^, p16^INK4A^, and p21^CIP1^ expression. **E.** Within disease cohorts, MaGE proxies tracked COVID-19 severity, and Crohn’s status and resolution. Figure created in BioRender: https://BioRender.com/5k0zctd

Upon selecting CpGs through mutual information, but before training predictors against gene expression, we conducted an enrichment analysis to validate the selection process, testing for overlap between the top MI-ranked CpGs and senescence-related traits and proteins in the EWAS Catalog ^44^. Following Benjamini-Hochberg (BH) correction, p14^ARF^ CpGs showed significant enrichment for smoking, age, cardiovascular conditions, alcohol, and mortality, while p21^CIP1^ CpGs overlapped significantly with smoking and age (Supp. Fig. 1). Both were also significantly enriched for CpGs associated with C-reactive protein (CRP). p14^ARF^ CpGs were additionally enriched for LTA and GZMA ^45–47^, which are involved in immune-mediated cytotoxicity and senescent cell clearance. In addition, p16^INK4A^ showed nominally significant associations with smoking, age and CRP.

To generate the MaGE predictors, samples were split into training (n=363) and held-out test (n=91) sets, and elastic net regression was applied to the top 200 mutual information-ranked CpGs for each target gene, after adjusting for age and surgical exposure. This yielded three individual MaGE predictors (MaGE-p14, MaGE-p16, MaGE-p21) plus a composite score (MaGE-composite), calculated as the arithmetic mean of the three individual predictors’ z-scores.

Predicted methylation-based expression correlated significantly with observed qPCR expression for all three genes in both training and held-out test data (training: Spearman’s ρ = 0.61, p14^ARF^; ρ = 0.53, p16^INK4A^; ρ = 0.41, p21^CIP1^; test: ρ = 0.57, p14^ARF^; ρ = 0.32, p16^INK4A^; ρ = 0.36, p21^CIP1^) (Supp. Fig. 2), confirming that whole-blood DNAm captures sufficient signal to proxy CD3+ T-cell-specific transcript abundance for each senescence marker.

### MaGE as a predictor of disease risk

To establish the biological interpretability and clinical relevance of MaGE proxies, we assessed their capacity to predict incident disease among 18,859 Generation Scotland participants across 10 years of follow-up, enabling its evaluation at a population scale. A total of 696 Cox proportional hazards regression models were run between the four predictors and 174 incident diseases (Supp. Data 1), considering diseases with a case threshold minimum of 30 incident diagnoses (median_case_ = 110.5, max_case_ = 1,527 for hypertension, see Supp. Data 2). To avoid over-adjustment for relevant mediators of the senescence phenotype, covariate adjustment was restricted to age and sex. Relatedness was regressed from the MaGE predictors prior to the Cox analyses. To investigate smoking (ever/never status) and sex differences, interaction models were computed (Supp. Data 3). Global MaGE values of all three senescence markers considered sex-specific bias (Supp. Fig. 3). An age-dependent divergence was observed, with males accumulating higher levels later on in life consistent with previous literature ^48^.

At nominal significance (P < 0.05) there were 219 associations across 70 unique diseases, 36 of which were shared across all three predictors (Fig. 2A). 45 Bonferroni-significant associations (P < 2.87 ×10^-4^) were observed between the three MaGE predictors and their composite for 18 unique incident diseases (Fig. 2B). Of those, 14, 12, 11 and 8 were observed for MaGE-composite and MaGE-p16, -p14 and -p21, respectively (Table 1). An R Shiny app aggregates the full suite of incident disease associations in a user-oriented manner (HTTPS://HSR-RSC-PROD3.MAYO.EDU:1919/MAGE_DISEASE_ASSOC_SEP2026/).

**Figure 2.**
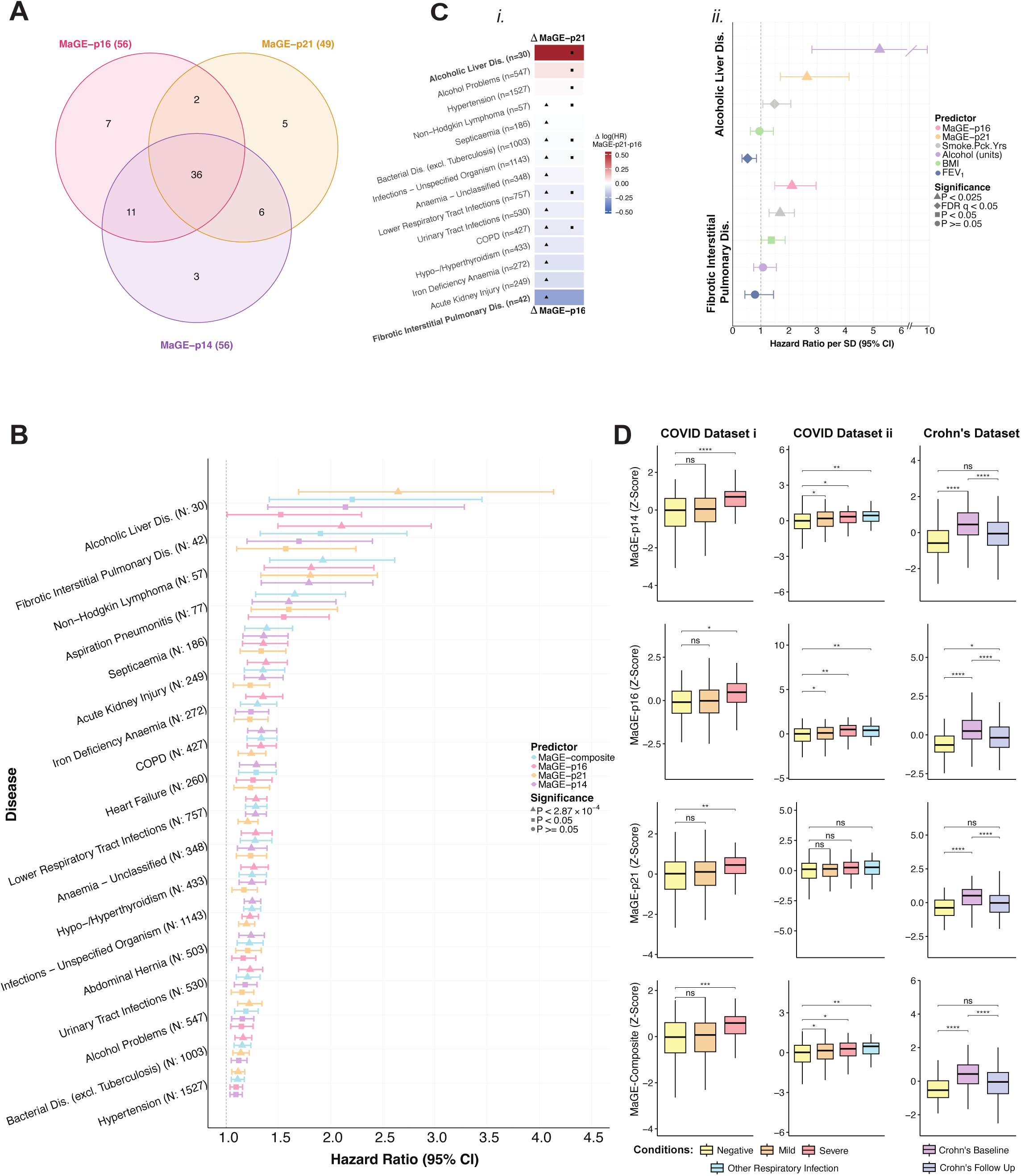
Senescence MaGE predictors associate strongly with disease onset and incidence. (**A**)Venn diagram of the 56 mutually significant diseases overlapped across all three proxies (P_nominal_ <0.05). MaGE-p16 in pink, -p21 in gold and -p14 in purple. (**B**) Forest plot of hazard ratios for 18 incident diseases with at least one Bonferroni-significant MaGE-disease pairing (P < 2.87 ×10^-4^). Results are arranged in ascending order of effect size. Significance is denoted by shape and proxy type by color (triangle: P _Bonferroni_ <2.87 ×10^-4^, square: P_nominal_ <0.05, circle:P_non-significant_ >0.05). MaGE-p16 is highlighted in pink, -p21 in gold, -p14 in purple and -composite in blue. Error bars indicate LCI and UCI, HR at 95% CI. (**C**) i. Difference (Δ) of the log (HR) of disease associations between MaGE-p21(Δ+, red) and MaGE-p16 (Δ-, blue), where at least one proxy is Bonferroni significant (P<2.87×10^-4^). ii. Forest plot of MaGE risk for Pulmonary Fibrosis and Alcoholic Liver Disease, compared with clinical factors (FEV_1,_ BMI, Smoking pack years, alcohol consumption, shown in: blue, green, grey and purple). Data are ordered by effect size, and significance is denoted by shape: triangle: P_Bonferroni_ < 0.025, diamond: BH-FDR q <0.05, square: P_nominal_ <0.05 and circle: P_non-significant_ >0.05. (**D**) MaGE predictors applied to three DNAm datasets comparing COVID-19 status (left panel - GSE179325; negative = 101, mild = 360, severe = 113, and middle panel - GSE167202; negative = 219, mild = 118, severe = 46 and other infection = 65) against controls, and Crohn’s disease status comparing pediatric cases at baseline (blood collected at diagnosis, n = 164), paired longitudinal follow-up (1–3 years post-diagnosis following intervention treatments, n = 164), and non-IBD controls (n = 74) (right panel - GSE112611). Group comparisons of predicted expression (Z-scores) were evaluated via linear regression adjusting for age and sex, followed by Bonferroni correction (*p < 0.05, **p < 0.01, ***p < 0.001, ****p < 0.0001).

**Table 1:** Overview of the 4 MaGE predictors and 14 epigenetic clocks^27,49–59^ considered as risk predictors.

|  |  |  |  |  | N Disease Associations (Cox Model)<br>Age and Sex Adjusted |  |
| --- | --- | --- | --- | --- | --- | --- |
| Predictor | Type | Outcome | Tissue | N CpGs in Clock | Benjamini-Hochberg: FDR<sub>q</sub> < 0.05 | Bonferroni : P < 0.05/174 |
| Hannum (2013) | First | Age (years) | Blood | 71 | 29 | 36 |
| Horvath (2013) | First | Age (years) | Multi-tissue | 353 | 17 | 27 |
| Lin (2016) | First | Age (years) | Blood | 100 | 12 | 17 |
| Zhang10 (2017) | Second | Mortality | Blood | 514 | 60 | 84 |
| PhenoAge (2018) | Second | Phenotypic age | Blood | 513 | 54 | 76 |
| Horvath Skin & Blood (2018) | First | Age (years) | Skin and blood | 391 | 28 | 40 |
| GrimAge v1 (2019) | Second | Mortality | Blood | 1030 | 68 | 102 |
| DNAmTL (2019) | NA | Telomere length | Blood, adipose | 146 | 47 | 60 |
| DunedinPoAm38 (2020) | Third/Pace | Pace of aging | Blood | 46 | 58 | 95 |
| DunedinPACE (2022) | Third/ Pace | Pace of aging | Blood | 173 | 72 | 103 |
| GrimAge v2 (2022) | Second | Mortality | Blood | 1030 | 72 | 105 |
| YingCausAge (2024) | First | Age (years) | Blood | 586 | 19 | 27 |
| YingDamAge (2024) | First | Age (years) | Blood | 1090 | 19 | 28 |
| YingAdaptAge (2024) | First | Age (years) | Blood | 1000 | 20 | 28 |
| MaGE-p14 (2026) | MaGE | p14<sup>ARF</sup> transcript abundance | Blood | 33 | 33 | 11 |
| MaGE-p16 (2026) | MaGE | p16<sup>INK4A</sup> transcript abundance | Blood | 17 | 39 | 12 |
| MaGE-p21 (2026) | MaGE | p21<sup>CIP1</sup> transcript abundance | Blood | 8 | 29 | 8 |
| MaGE-composite (2026) | Arithmetic mean | NA | Blood | NA | 38 | 14 |

### MaGE recapitulates previously described senotype-associated organ specificity

MaGE-disease association pairings were largely distinct, in line with prior experimental evidence for tissue-preferential roles of p21^CIP1^ and p16^INK4A^ to liver and lung, respectively (Fig. 2C outlines this specificity by the difference in log effect sizes of MaGE-p16 and -p21, extended to all three proxies in Supp. Fig. 4). The study’s strongest hazard ratio was for MaGE-p21 was with Alcoholic Liver Disease (N=30, HR = 2.64, 95% CI [1.693, 4.138], P = 1.94×10^-5^) (Fig. 2B). MaGE-p21 was also associated with the 10-year onset of Alcohol Problems (N=547) (N=547, HR = 1.22 [1.11, 1.34], P = 4.13×10⁻ ^5^). At FDR q<0.05 MaGE-p21 was additionally associated with incident fatty liver disease (HR 1.55 [1.18–2.03], P=1.7×10⁻ ^3^). In contrast, MaGE-p16 showed its strongest association with Interstitial Pulmonary Fibrosis (N=42, HR = 2.10 [1.49, 2.96], P = 1.99×10⁻ ^5^) (Fig. 2C). At FDR q<0.05 MaGE-p16 was also associated with incident asthma (HR 1.20 [1.08–1.32], P = 5.1×10⁻ ^4^) (Fig. 2C). Together, these associations suggest distinct disease patterns for MaGE-p21 and MaGE-p16, highlighting known preferential associations with hepatic and pulmonary outcomes, respectively.

Four diseases showed concordant, Bonferroni-significant associations across all four predictors, including Lower Respiratory Tract Infections (HR MaGE-p16=1.28 [1.18, 1.38], P=3.88×10⁻ ^10^).

### MaGE associations are comparable in magnitude to established clinical risk factors

To contextualize the magnitude of our two strongest associations, we benchmarked MaGE-p16 and MaGE-p21 against established clinical risk factors (FEV*₁*, Smoking Pack Years, Alcohol Consumption, BMI), adjusted for age, sex, and (for FEV*₁*) height (Fig. 2C). Of the five predictors considered for IPF, MaGE-p16 showed a stronger association than Smoking Pack Years, the only clinical measure to survive Bonferroni correction (HR _MaGE-p16_ =2.10, P=1.99×10⁻ ^5^; HR_Smoking Pack Years_= 1.68 [1.29, 2.19], P=1×10⁻ ^4^). Similarly, for ALD, only MaGE-p21 and Alcohol Consumption remained Bonferroni-significant (HR _MaGE-p21_=2.64, P=1.93×10⁻ ^5^; HR _Alcohol Consumption_= 5.22 [2.88, 9.68], P=1.5×10⁻ ^7^). Together, these findings indicate that MaGE proxies may capture disease risk on par with, or exceeding, well-established clinical risk factors.

To determine whether these associations persisted after adjustment, we tested MaGE-p16 and MaGE-p21 in a fully adjusted model incorporating age, sex, BMI, FEV*₁*, Smoking Pack Years, and Alcohol Consumption. MaGE-p16 remained significantly associated with IPF onset (N=28, HR=1.73 [1.14, 2.63], P=0.011), while MaGE-p21 remained significantly associated with ALD onset (N=28, HR=2.51 [1.55, 4.05], P=1.8×10⁻ ^4^), indicating that both associations persist after adjustment for established clinical risk factors.

### Validation in disease cohorts track COVID-19 severity and Crohn’s resolution via intervention

Having established MaGE’s associations with incident diseases in Generation Scotland, we next examined MaGE in independent cohorts spanning acute and chronic inflammatory disease states (Fig. 2E). In two independent COVID-19 datasets ^60,61^, all four predictors were significantly elevated in severe (hospitalized) cases relative to negative controls (Bonferroni-adjusted P<0.05, age- and sex-adjusted), with the exception of p21^CIP1^ in the second dataset. In a pediatric Crohn’s disease dataset ^62^, newly diagnosed cases showed markedly higher predicted senescence expression than controls (Bonferroni-adjusted P<0.001), which normalized to control levels following 1–3 years of anti-inflammatory treatments, consistent with a treatment-associated resolution of inflammation. Consistent observations were observed in Generation Scotland, where incident Crohn’s disease (N=40, HR _MaGE-p16_=1.76 [1.24, 2.48], P=1.4×10⁻ ^3^) and coeliac disease (N=60, HR _MaGE-p16_=1.59 [1.21, 2.11], P=1.0×10⁻ ^3^) were significantly associated with MaGE-p16 (BH-adjustment), alongside further associations for coeliac disease with MaGE-p14 and MaGE-composite. Incident COVID-19, however, was not available for our analysis. Together, these findings support the generalizability of MaGE as a blood-based indicator of senescence-associated biology across diverse disease settings and suggest that MaGE may be responsive to changes in disease state.

### MaGE shows disease-association profiles distinct from established epigenetic clocks

To contextualize MaGE associations with incident disease, we compared the four MaGE proxies with 14 established epigenetic clocks ^27,49–59^ previously evaluated in Generation Scotland ^31^. Clock-disease Cox regression models have been previously described ^31^. Briefly, associations were assessed for the same set of 174 incident diseases, over 10 years of follow-up, with adjustment for age and sex (full list in Supp. Data 4).

We first assessed whether the MaGE proxies achieved comparable predictive performance to a panel of 14 established clocks. There was a total of 390 Bonferroni-significant associations, with the majority attributed to third- and second-generation clocks which had on average 53.5 and 48.2 associations per predictor, followed by MaGE proxies (11.2 per proxy) and first-generation clocks (3.3).

Among nominally significant disease associations (P<0.05, 149 diseases), the mean absolute log-hazard ratio for MaGE proxies was approximately ∼71% greater in magnitude than for first generations clocks, whereas MaGE and second-generation clocks showed more similar effect-size distributions – with the latter marginally larger ∼8%.

To determine whether this performance reflected biological signal distinct from existing clocks rather than redundancy, we examined the structure of predictor effect-size profiles. Principal component analysis (PCA) predictor effect-size profiles (Fig. 3B) revealed distinct separation of the four MaGE proxies along PC2, whereas clocks clustered by generation along PC1 (PC1, 56%; PC2, 16.3%).

**Figure 3.**
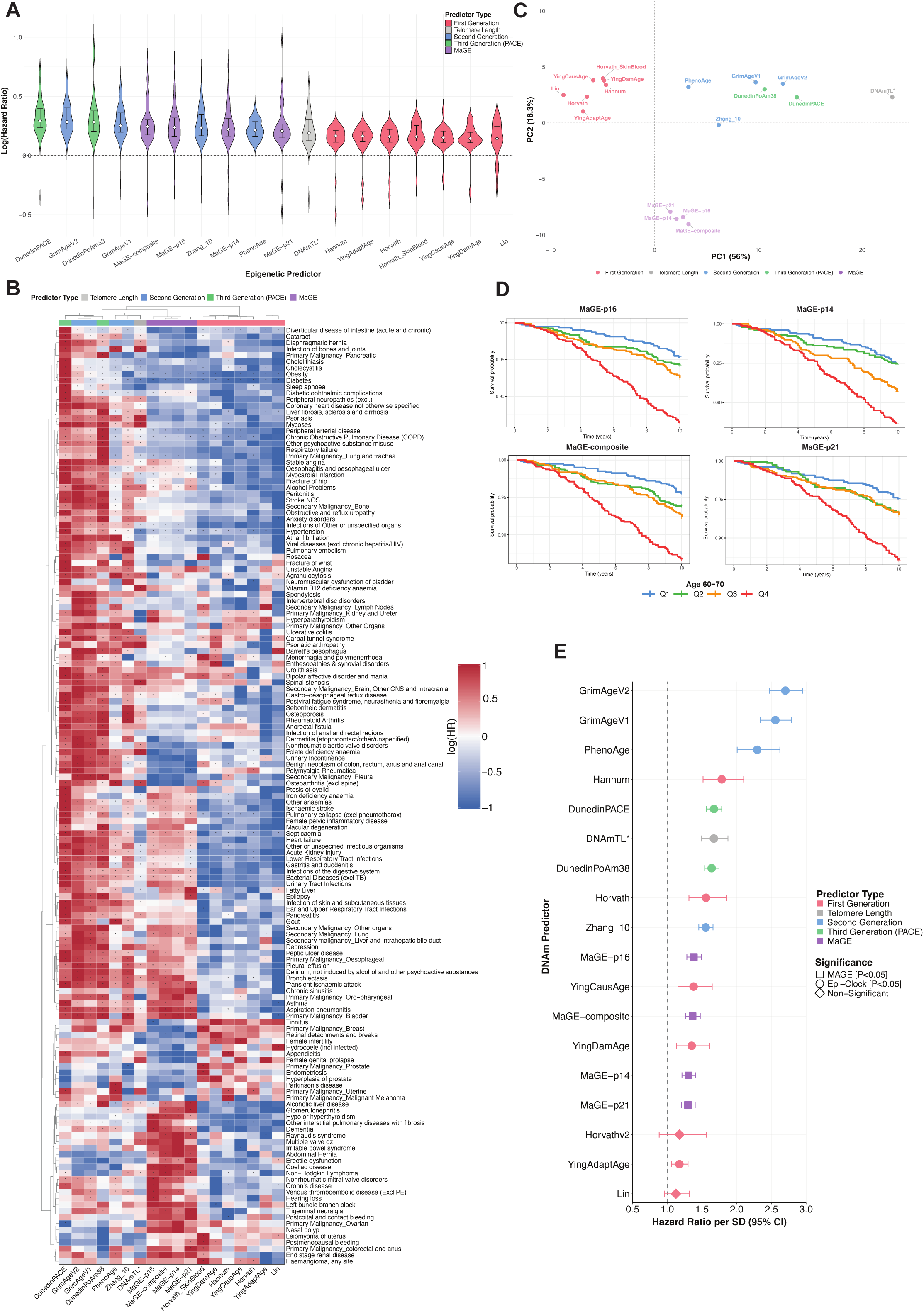
Senescence MaGE predictors are comparable to 2^nd^ and 3^rd^ generation epigenetic clocks for disease associations yet offer a complementary disease signal and associate strongly with all-cause mortality. (**A**) Effect size distribution of the 149 nominally significant (P< 0.05) associations across all four MaGE predictors and fourteen, cross-generational epigenetic clocks. Ordered by descending median effect size shown as violin plots with interquartile range. First-generation clocks are highlighted in salmon, telomere length in gray, second-generation in blue, third/pace in green and MaGE proxies in purple. *Telomere length HR effect sizes have been multiplied by -1 for visual display purposes and uniformity across the following panels: (A, B, C, D). (**B**) Principal component analysis (PCA) of 149 nominal (P <0.05) predictor effect-size profiles of all four MaGE predictors (purple) and fourteen, cross-generational epigenetic clocks (colored by generation as per Fig. 3A). (**C**) Hierarchical clustering of log-transformed and row-normalized effect sizes across all 149 significant associations (P<0.05). Marked tiles (*) indicate Bonferroni significance (P<2.87×10^-4^). Range is depicted as follows: negative (min) and positive (max) denoted by color intensity, blue for negative (indicating smaller relative effect sizes) and red for the later. (**D**) Kaplan-Meier curves stratified by senescence-proxy quartile (N= 251, age range: 60-70), where MaGE-quartiles are separated by color (blue for Q1, green for Q2, orange for Q3 and red for Q4). (**E**) Survival models compared effect sizes for 10-year all-cause mortality of both MaGE and fourteen epigenetic clocks. Models were adjusted for age and sex (N_cases_ = 842). Shape portrays significance and predictor type, square for significant epi-clock associations and a circle for significant MaGE associations (P <0.05, no multiple testing required). Non-significance is denoted by a diamond shape (P >0.05). Predictors are colored as previous.

Consistent with the PCA, this separation persisted after applying unsupervised hierarchical clustering to predictor disease-association profiles. Hierarchical clustering of effect sizes across all nominal associations (Fig. 3C) grouped the four MaGE proxies separately from the epigenetic clocks, which clustered broadly by generation.

In an orthogonal analysis we tested the degree of overlap between the MaGE proxies and the residual age-acceleration of 14 published epigenetic clocks ^27,49–59^ (Supp. Fig. 5). MaGE proxies clustered closely amongst themselves, distinct from epigenetic clocks, correlating most strongly with telomere length (DNAmTL) (shorter predicted telomere length corresponding to higher MaGE output).

Collectively, these analyses indicate that MaGE proxies have a pattern of disease associations that differs from those captured by established epigenetic clocks, whilst exceeding the broad-range predictive performance of first-generation clocks for incident disease.

We next assessed whether MaGE associations extend to all-cause mortality. Kaplan-Meier analysis in the 60-70 age bracket (selected to maximize mortality event numbers, N = 251) revealed progressively lower survival with increasing senescence-proxy quartile (Fig. 3D), a trend generally consistent across other age groups (Supp. Fig. 6). Ten-year survival probabilities were closely clustered across the lower three quartiles for all four MaGE predictors but showed a markedly larger decrement in the highest quartile, indicating that the reduction in survival was concentrated in individuals with the highest MaGE levels. This separation was greatest for MaGE-p16, which showed the lowest survival probability of any proxy with an absolute reduction of 0.09 (Q1_MaGE-p16_: 0.96 - Q4_MaGE-p16_: 0.87).

To contextualize the magnitude of this mortality association, we compared MaGE and clock effect sizes directly within the same survival framework. All four MaGE proxies were associated with 10-year all-cause mortality (N_cases_ = 842), with MaGE-p16 showing the largest association (HR_MaGE-p16_ = 1.38 per SD, 95% CI [1.28–1.49], P = 2.7×10⁻ ¹⁷). The magnitude of association for MaGE-p16 fell within the range observed for first-generation clocks but was smaller than that of second- and third-generation measures, developed to capture mortality and health-related outcomes (Fig. 3E). MaGE proxies remained significantly associated with mortality after further adjustment with GrimAge (V1 and V2), indicating an independent contribution to mortality risk (Supp. Data 5).

Together, these findings position MaGE as a DNAm-derived measure associated with both incident disease and mortality, while exhibiting disease-association profiles distinct from epigenetic clocks.

### Genome-wide association analyses reveal loci associated with senescence, inflammation, and disease

We next investigated the genetic architecture of MaGE using genotype data from Generation Scotland (Fig. 4A). Genome-wide association analyses identified loci shared across the three MaGE predictors as well as predictor-specific associations. Shared loci included variants mapping to NFKB1 and LTβR. Predictor-specific associations included CSF3, which associated with MaGE-p21, and CHFR, which associated with MaGE-p16. Cross-referencing lead variants with the NHGRI-EBI GWAS Catalog identified previous associations with hematologic traits, inflammatory disease, type 1 diabetes, cancer, and circulating CRP levels.

**Figure 4.**
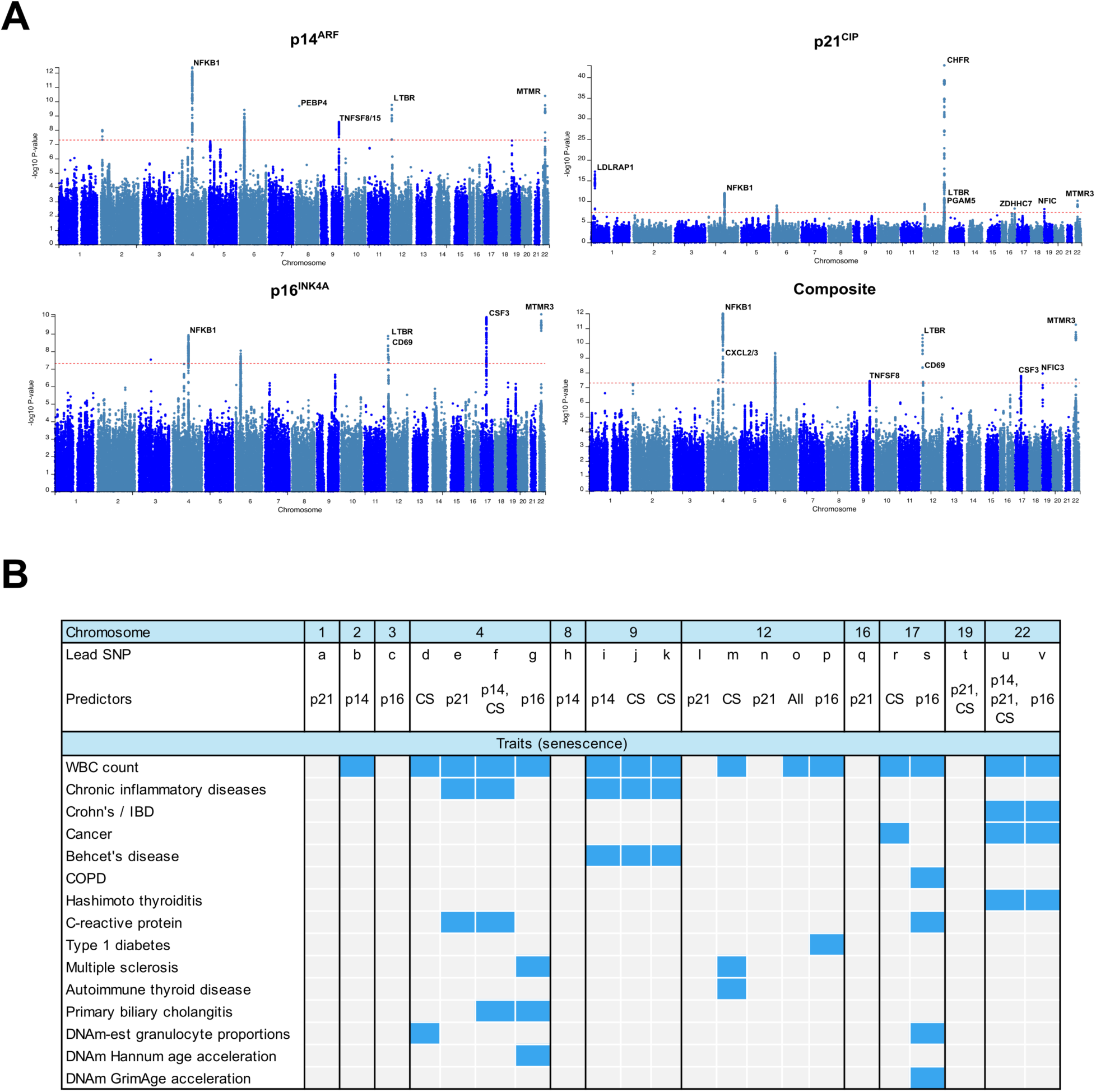
Senescence- and disease-related genomic loci associate strongly with senescence MaGE predictors. (**A**) Genome-wide association analysis (GWAS) of MaGE-p14, MaGE-p16, MaGE-p21 and MaGE-Composite. Red dotted line is the minimum significance threshold for MaGE-associated SNPs (P < 5×10^-8^). (**B**) NHGRI-EBI GWAS Catalog entries for all lead SNPs for all MaGE predictors (blue = hit, grey = no hit). Lead SNPs are denoted by letters; full coordinates are provided in Supp. Data 6.

## Discussion

### Overview

We developed Methylation-associated Gene Expression (MaGE) predictors to estimate expression of three senescence-associated transcripts, p16^INK4A^, p14^ARF^ and p21^CIP1^, from whole-blood DNAm. These measures were prospectively associated with broad morbidity and all-cause mortality, showing distinct patterns of association across the individual predictors. MaGE exhibits a different pattern of disease associations and individual-level correlation structure to epigenetic aging clocks. Genetic analyses identified loci mapping to pathways implicated in senescence, inflammation, and disease. Together, these findings support MaGE as a scalable approach for studying senescence-associated molecular phenotypes in human populations. Importantly, they enable retrospective application to existing population cohorts and biobanks with archived blood DNAm.

### Disease associations are consistent with marker-specific senescence biology

The distinct disease-association patterns observed for the individual MaGE predictors are consistent with emerging evidence that p16^INK4A^- and p21^CIP1^-associated senotypes are biologically distinct. scRNA-seq has identified differences in the secretory profiles of p16^INK4A^-high and p21^CIP1^-high populations across human and murine tissues ^63^, while experimental clearance of these populations has produced divergent effects on lifespan, osteoporosis and fracture healing^12,64–66^. In Generation Scotland, the strongest association for MaGE-p21 was with ALD, consistent with prior links between p21^CIP1^ and ALD ^67^ and hepatic fibrosis ^68,69^. Conversely, the strongest MaGE-p16 association was with IPF, consistent with previous studies linking p16^INK4A^ to pulmonary disease severity ^70^ and survival ^71^. With two functionally distinct markers resolving prospectively onto their expected target organs, this demonstrates that MaGE captures marker-specific and grounded senescence biology rather than a generic aging signal, and points toward organ-directed risk stratification.

### Mortality & Clinical Benchmark

Higher MaGE scores were associated with lower 10-year survival, with the greatest separation observed in the highest quartile. All four MaGE measures were also associated with all-cause mortality in age- and sex-adjusted models, consistent with our previous observation that circulating biomarkers associated with cellular senescence are related to mortality in humans ^72^. Notably, the highest quartile of MaGE-p16 exhibited the lowest survival probability of any of the senescence gene proxies (Fig. 3D)., consistent with a phase 2 senolytic intervention trial, where skeletal benefit was confined to the highest tertile of CD3+ T-cell p16^INK4A^ expression ^73^. When benchmarked against established epigenetic clocks, MaGE showed mortality associations within the range of first-generation clocks, whereas second- and third-generation measures showed stronger associations with mortality. This difference is not unexpected, as several later-generation clocks were explicitly trained on mortality, morbidity or related health phenotypes.

### Genetic validation suggests MaGE captures in vivo senescence pathways

Our genome-wide analyses provide orthogonal evidence that senescence MaGE captures core senescence biology. The leading hit across all three independent MaGE predictors was *NFKB1*, a core component of the NF-κB signaling that drives the SASP and whose loss accelerates cellular senescence and premature aging in mice^74^. Its upstream activator *LTβR* was recovered alongside it ^75^. The recovery of a shared NF-κB axis alongside suggests that MaGE reflects senescence biology in vivo.

### Potential applications of MaGE in clinical research

An important potential application of MaGE is in the development and evaluation of senotherapeutic interventions where MaGE could facilitate participant stratification. In our phase 2 trial of dasatinib plus quercetin, skeletal benefit was concentrated solely among women with the highest baseline CD3+ T-cell p16^INK4A^ expression ^73^. This finding raises the possibility that a scalable DNAm-derived proxy could facilitate participant stratification in future senotherapeutic trials, although whether MaGE identifies treatment-responsive populations similarly will require prospective evaluation. Beyond stratification, MaGE could have value as a pharmacodynamic reponse biomarker in early-phase and dose-ranging studies, providing a much earlier indication of biological activity than downstream clinical outcomes. The reduction of elevated MaGE following anti-inflammatory treatment in paediatric Crohn’s disease suggests that MaGE can change with disease or treatment state, yet whether a similar response will replicate to senotheraputic intervention remains unknown. Prospective studies incorporating serial MaGE measurements alongside direct measures of senescent cell burden will therefore be needed to establish its utility for dose selection and treatment-response assessment.

### Study Limitations

Several limitations should be considered when interpreting these findings. First, MaGE was trained in a relatively small cohort of older women. Reassuringly, no significant difference in qPCR expression of p14^ARF^, p16^INK4A^ or p21^CIP1^ was observed between ovariectomy groups (Supp. Fig. 7). In addition, we report an absence of significant findings in disease interaction models considering sex-stratified analyses.

MaGE uses whole-blood DNAm to approximate expression of senescence-associated transcripts measured in isolated CD3+ T cells. Thus, although the transcriptional phenotype itself is cell-specific, the extent to which MaGE reflects CD3+ T-cell-intrinsic biology versus systemic factors captured by whole-blood DNAm remains to be determined. Independent validation, direct comparison with cellular senescence measures, and prospective studies incorporating serial sampling will be important next steps toward establishing the biological and clinical utility of MaGE.

Although MaGE was prospectively associated with multiple incident diseases, these observational associations do not establish causality or clinical predictive utility, and several of the strongest disease-specific findings, including those for ALD and IPF, were based on relatively small numbers of incident events and warrant replication. Similarly, associations observed in independent disease cohorts support generalizability across clinical settings but cannot resolve the contributions of cellular senescence and closely interconnected processes such as inflammation to the MaGE signal.

## Methods

### MaGE Predictor Creation

#### CD3+ T-cell isolation and qPCR

CD3+ T-cells were enriched directly from whole blood using AutoMACS™ pro separator (Miltenyi Biotec, Gaithersburg, MD) after labeling with CD3 magnetic microbeads (Miltenyi Biotec) following the manufacturer’s protocol. RNA was isolated from isolated cells using Trizol Reagent (Invitrogen, Austin, TX, USA) following the manufacturer’s protocol. RNA was then quantified using a NanoDrop 8000 spectrophotometer (ThermoFisher, Waltham, MA, USA). cDNA was synthesized from 1µg of total RNA using M-MLV reverse transcriptase (Invitrogen) and real-time PCR was performed with PerfeCTa FastMix II Low ROX (QuantaBio) and the QuantStudio 5 Real-Time PCR system (Thermo Fisher). Gene expression was analyzed by the ΔΔCT method and normalized to the reference gene TBP.

#### MOA-2 cohort study Data preparation

To facilitate cross-cohort validation, CpG sites were first restricted to those present in both the Mayo Clinic Cohort Study of Oophorectomy and Aging-2 (MOA-2) cohort ^43^ and the Generation Scotland (GS) ^42^ dataset. Missing methylation values were imputed using per-CpG means calculated exclusively from the training samples, and these means were then applied to impute missing values in the test set. CpG sites with all-missing or zero-sum methylation values across samples were removed. A single global train/test split was performed prior to any preprocessing or model fitting (Supp. Table 1), allocating 80% of samples to training (n = 363) and 20% to held-out testing (n = 91). Stratification by prior oophorectomy status (Bilateral Salpingo-oophorectomy (BSO), yes/no) was applied to ensure balanced representation across splits.

#### Target Variable Transformation and Covariate Adjustment

To adjust for covariates before creating the predictors, a linear regression model was fit on the training samples, with the log-transformed target as the response and age at visit and oophorectomy status as predictors. Samples with missing values in the target variable (qPCR gene expression) were excluded on a per-variable basis. Residuals from this model were used as the outcome for model training. The same regression model (fit on training data only) was used to compute residuals for the test set. The adjusted residuals were then standardized to zero mean and unit variance (using training statistics).

#### Feature Selection

To reduce the number of features prior to model fitting, mutual information (MI) between each CpG site and the adjusted, standardized target variable was estimated on training samples only. CpG features were first discretized into 15 quantile-based bins using KBinsDiscretizer from scikit-learn, and MI scores were computed using mutual_info_regression. The top 200 CpG sites by MI score were selected as features for each target variable. The selected CpGs were z-score standardized (mean = 0, std = 1) using per-CpG statistics computed from the training data.

#### Elastic Net Regression

An elastic net regression model was fit for each target variable using ElasticNetCV from scikit-learn, with an L1 ratio of 0.5 (equal weighting of L1 and L2 penalties) and 5-fold cross-validation for regularization parameter (α) selection. The resulting Methylation-associated Gene Expression (MaGE) predictors performances were evaluated on both the training and held-out test sets using Spearman’s rank correlation between observed and predicted values (Supp. Fig. 2).

### EWAS Catalogue Enrichment Analysis

To evaluate the biological relevance of CpG sites with the highest mutual information (MI) relative to expression of p14, p16 and p21, enrichment analyses were conducted of the top 1000 MI ranked CpGs against the EWAS Catalog ^44^. Since a significant proportion (∼36%) of the Catalog’s unique study identifiers (Study IDs) included 450K studies and the MI-expression associated CpGs were derived from EPIC v1 data, we restricted our top 1000 CpGs for each senescence-associated protein to CpGs present on both the EPIC and 450K array. This resulted in 434, 515, and 503 CpGs for p14^ARF^, p16^INK4A^ and p21^CIP1^ respectively.

We excluded EWAS Catalog entries with missing effect size estimates and limited our search to blood-derived tissues (whole blood, PBMC, and CD4+ T-cells). Associations were partitioned into two categories: phenotypic traits and protein biomarkers. Phenotypic traits were harmonized and filtered for 11 categories with established links to senescence: aging, mortality, smoking, cancer, Alzheimer’s disease, chronic obstructive pulmonary disease, cardiovascular disease, rheumatoid arthritis, kidney disease, obesity, and alcohol consumption. For protein biomarkers, a curated panel of 108 senescence-associated proteins was assembled, encompassing components of SASP, inflammaging mediators, immune surveillance effectors, and immune checkpoint molecules. Over-representation of predictor CpGs among trait-associated CpGs was assessed using one-sided Fisher’s exact tests, using 450K array CpGs present on the EPIC array as the background set. Resulting p-values were adjusted for multiple comparisons using the Benjamini-Hochberg procedure.

### External Validation of MaGE Predictors in Disease Cohorts

To evaluate the ability of MaGE predictors to detect senescence burden in disease states, we applied the trained models to three independent, publicly available DNA methylation datasets from the Gene Expression Omnibus (GEO):

- GSE179325 - Whole blood Illumina EPIC v1 methylation data from COVID-19 patients and healthy controls ^60^. Subjects were classified as Negative, Mild, or Severe based on disease status. Phenotype covariates included age and sex.
- GSE167202 - Whole blood Illumina EPIC methylation data from a second COVID-19 cohort ^61^. Subjects were categorized by severity score (1–4, collapsed into Mild [1–2] and Severe [3–4]), with additional groups for COVID-negative controls and patients with other infections. ICU-admitted negative controls were excluded to avoid confounding intensive-care-associated methylation changes.
- GSE112611 - Whole blood Illumina EPIC methylation data from pediatric Crohn’s disease patients and healthy controls ^62^. Crohn’s patients were sampled at baseline and follow-up, yielding three groups: Control Baseline, Crohn’s Baseline, and Crohn’s Follow-Up.

#### Data Processing

For each dataset, missing beta values were imputed using per-CpG row-mean imputation. Methylation matrices were then row-scaled (Z-score normalization across samples for each CpG site) prior to model application, consistent with the normalization applied during model training.

#### Prediction of Senescence Marker Expression

For applying each MaGE predictor (MaGE-p14, MaGE-p16 and MaGE-p21) to methylation datasets, the model intercept and CpG coefficients were loaded from the trained elastic net model weights. Predictions were calculated as the sum of the intercept and the weighted methylation values for each CpG site. CpGs present in the model but absent from a given dataset were excluded, and predictions were computed using the remaining available CpGs.

Raw prediction values were Z-score transformed across all samples within each dataset and predictor to enable cross-predictor comparability. A composite senescence score (MaGE-composite) was calculated as the arithmetic mean of the three individual MaGE Z-scored predictions (MaGE-p14, MaGE-p16 and MaGE-p21) for each sample.

#### Statistical Analysis of Group Differences

To assess differences in predicted senescence marker expression between disease states in the disease cohort analysis (Fig. 2D), linear regression models were fitted with predicted expression (Z-score) as the outcome and disease group as the primary independent variable, adjusting for age and sex as covariates. Pairwise contrasts between negative states versus other disease stages were extracted from each model. As our aim was to characterize the univariate association of each MaGE predictor (including MaGE-composite) with disease status (rather than to compare relative performance or effect sizes across predictors), Bonferroni correction was applied within each predictor across its disease-stage comparisons. Hence, each predictor was treated as an independent test family, following the general principle of per-predictor correction over cross-predictor pooling used in large-scale clock-disease association studies ^31^. Results were visualized as boxplots with Bonferroni-adjusted significance brackets using the ggpubr R package.

### Generation Scotland: Disease Risk Analysis

#### Generation Scotland cohort

Generation Scotland is a family-based cohort of 24,084 volunteers recruited 2006– 2011, (age at baseline: 17-99 years) described in full ^76^. All individuals were invited to complete questionnaires and to attend a baseline clinic visit for further questionnaires and testing, resulting in a detailed collection of lifestyle, clinical, health, cognitive and sociodemographic data. The majority also consented for biosamples collection, including blood, and provided consent for researchers to access their medical records via data linkage to primary and secondary care codes ^76^.

#### DNA methylation profiling

As previously described, DNAm was quantified on the Illumina EPIC 850K array (851,610 CpGs post-QC) for 18,859 volunteers after 10 withdrawals, as well documented in previous published literature^31,76^. We considered the same epigenetic clocks as Mavrommatis et al. 2025 ^31^. In brief, age acceleration estimates for the fourteen clocks was computed within the Biolearn platform^77^ followed by a regression on age and relatedness.

#### Disease coding, mortality and covariates

Incident disease outcomes were defined from linked primary and secondary care records using the consensus definitions of Kuan et al. 2019 and derived exactly as in Mavrommatis et al. 2025 (first diagnosis, prevalent cases excluded, ≥30 incident cases over 10-year follow-up, giving 174 outcomes) ^31,78^. A comprehensive list of 174 diseases observed and a number of incident events (events occurring after blood draw) along with mean age at diagnosis – utilized for both MaGE and Clock analyses can be found in Supplementary Data 2. Primary care records were available for around 40% of participants. Mortality was ascertained via Community Health Index linkage to National Records of Scotland and formed part of the censoring criteria (follow-up to April 2022)^31^.

We considered demographic, lifestyle and health variables as covariates in the regression models with each MaGE/Clock and disease outcome. The primary analysis, relating MaGE with disease, used basic models adjusted for age and sex only, as computed and considered in Mavrommatis et al. 2025 – ensuring harmonization of MaGE/Clock models. Likewise, survival analysis also received basic adjustment. The clinical-benchmark models used two additional, MaGE-specific covariate setups: (i) models with FEV_1_ as the primary predictor were additionally adjusted for standing height; and (ii) fully adjusted clinical models controlled simultaneously for FEV_1_, BMI, smoking pack years, and alcohol consumption (units consumed in the week prior to the blood draw/baseline questionnaires), alongside age and sex. FEV_1_ was measured by spirometry at the baseline clinic visit. Because covariate data collection differs across these variables, case numbers vary between models considered as part of the clinical analysis; detailed lists can be found in Supplementary Data 7.

### Association between epigenetic predictors and disease outcomes

#### MaGE

Cox proportional hazards regression (survival package, *coxph()* R version 3.8.6) related each of the four MaGE predictors to 10-year onset of the 174 incident diseases (≥30 cases), giving 696 models. Models adjusted for age and sex only; to avoid over-adjustment for upstream mediators of the senescence phenotype, no further covariates were included. Relatedness (modelled as a kinship matrix) was regressed from the MaGE predictors prior to the Cox analyses, via the lmekin function of the coxme R package (v.2.2-22) ^79^; residuals from these models were taken forward for the main analyses as in our previous work^31^. Age and estimated white-blood-cell proportions were not regressed out of the MaGE predictors beforehand: the MaGE models were trained on target variables that had already been adjusted for age during predictor training, and was further adjusted in the Cox models, making additional regression redundant. Sex and smoking (ever/never) interaction models were run as previously described (pack years excluded when stratifying or testing interactions) ^31^. The proportional-hazards assumption was assessed via Schoenfeld residuals (cox.zph) and was met (Supp. Data 1). Multiple-testing correction used a Bonferroni threshold of P < 0.05/174 = 2.87×10⁻ ^4^ for disease outcomes and P < 0.05 for the single all-cause mortality test per predictor. Benjamini–Hochberg FDR-adjusted results (q < 0.05) are reported alongside. The composite predictor (MaGE-Composite) was the arithmetic mean of the three individual predictors’ z-scores.

Survival analyses were conducted via Cox model regression, with identical parameters to basic disease models. Kaplan Meier curves were generated via the *survfit* function of survival package v. 3.8.5.

#### MaGE versus clinical risk factors

MaGE-p16 and MaGE-p21 were benchmarked against FEV1, smoking pack years, alcohol consumption, and BMI for IPF and ALD, respectively. Each predictor was related to disease onset adjusting for age and sex (and, for FEV1, height), then in the fully adjusted clinical model above. As two diseases were tested, the Bonferroni threshold was P < 0.05/2 = 0.025.

#### MaGE versus epigenetic clocks

The 14 epigenetic clocks were related to the same 174 diseases in the identical Generation Scotland dataset under near-identical parameters, as described previously^31^. Minor differences in case counts arise from census-coding revisions for a small number of non-significant outcomes (Supp. Data 8).

### GWAS Functional Mapping and Annotation

#### GWAS

The GWAS was performed using the same dataset and similar methods as outlined in Dabriowski et al. ^80^. Briefly, genotype–phenotype association analyses were performed using a linear mixed model GWAS implemented in fastGWA GCTA^81^. A sparse GRM approach was used to adjust for sample-relatedness. Overall, 15,871 overlapping individuals with non-missing genotype and phenotype data were included from Generation Scotland. Variants with MAF < 0.01 or a missingness rate > 0.1 were excluded from the analysis. All coordinates were based on human reference genome assembly GRCh37/hg19 (http://www.ncbi.nlm.nih.gov/assembly/2758/).

#### FUMA analysis

Summary statistics from the four MaGE predictor GWAS were processed using the FUMA (Functional Mapping and Annotation of Genetic Associations) platform. To identify genomic risk loci, independent significant SNPs were defined based on a default threshold (p < 5×10^-8^) and local linkage disequilibrium (LD) structure (within a 250kb window).

The following parameters were set for lead SNPs and candidate SNPs identification:

- Sample size (N): 15871
- Maximum P-value of lead SNPs (<): 5e-8
- Maximum P-value cutoff (<): 0.05
- R^2^ threshold to define lead SNP (> or =): 0.6
- 2^nd^ R^2^ threshold to define lead SNPs (> or =): 0.1
- Reference panel population: UKB release2b 10k White British
- Include variants in reference panel (non-GWAS tagged SNPs in LD): Yes
- Minimum minor allele frequency: 0
- Max distance between LD blocks to merge into a locus: 250 kb

Default settings were kept for the following: 1. Gene Mapping (positional mapping); 2. Gene Mapping (eOTL mapping), 3. Gene Mapping (3D Chromatin Interaction mapping), 4. Gene types, 5. MHC region, and 6. MAGMA analysis. Using lead SNP rsIDs, we searched the GWAS catalog within FUMA for traits associated with lead SNPs (Fig. 4B, see Supp. Data 6 for SNP rsIDs).

#### Gene Prioritization for Visualization

To identify the most likely effector genes for Manhattan plot annotation, a dual-prioritization strategy was employed. First, MAGMA gene-based association scores were calculated to provide an aggregate statistical signal for each locus. Second, because genomic risk loci frequently encompass multiple genes in high LD, we performed functional prioritization. MAGMA genes were selected for labeling SNPs based on their established roles in cellular senescence, cell-cycle checkpoint regulation (e.g., CHFR, POLE), SASP (e.g., CSF3, CXCL), or mitochondrial homeostasis (e.g., PGAM5). See Supplemental Data 6 for full list of MAGMA genes and SNPs, as well as female-only analysis SNPs.

### Ethical Considerations, Acknowledgments and Data availability

#### Ethics and consent

Ethics, consent and Research Tissue Bank status for Generation Scotland are as described previously ^31^ (NHS Tayside REC 05/S1401/89; RT B 15/0040/ES). All participants provided broad written informed consent and the study conformed to the Declaration of Helsinki. Self-reported sex was cross-referenced against genetic data. Mayo Clinic Cohort Study of Oophorectomy and Aging-2 (MOA-2) has received approval by both Mayo Clinic and Olmsted Medical Center Institutional Review Boards (number 18-008476) and all participants provided written and informed consent ^82^.

## Data Availability

The source data that were analyzed during the current study are not publicly available due to them containing information that could compromise participant consent and confidentiality. Data can be obtained from the data owners. Instructions for accessing Generation Scotland data can be found here: (https://www.ed.ac.uk/generation-scotland/for-researchers/access); the GS Access Request Form can be downloaded from this site. Completed request forms must be sent to to be approved by the Generation Scotland Access Committee.
Mayo Clinic Cohort Study of Oophorectomy and Aging-2 is currently not publicly available but can be made available upon request.
External validation datasets are publicly available from GEO under accessions GSE179325, GSE167202 and GSE112611.

https://www.ncbi.nlm.nih.gov/geo/query/acc.cgi?acc=GSE179325

https://www.ncbi.nlm.nih.gov/geo/query/acc.cgi?acc=GSE167202

https://www.ncbi.nlm.nih.gov/geo/query/acc.cgi?acc=GSE112611

https://github.com/dsimps1993/Methylation-associated-Gene-Expression-Predictors/

## Data availability

The source data that were analyzed during the current study are not publicly available due to them containing information that could compromise participant consent and confidentiality. Data can be obtained from the data owners. Instructions for accessing Generation Scotland data can be found here: (https://www.ed.ac.uk/generation-scotland/for-researchers/access); the ‘GS Access Request Form’ can be downloaded from this site. Completed request forms must be sent to to be approved by the Generation Scotland Access Committee.

Mayo Clinic Cohort Study of Oophorectomy and Aging-2 is currently not publicly available but can be made available upon request.

External validation datasets are publicly available from GEO under accessions GSE179325, GSE167202 and GSE112611. Code for analysis available at https://github.com/dsimps1993/Methylation-associated-Gene-Expression-Predictors/blob/main/scripts/Disease_Datasets_MaGE_BF_081826.R

## R Shiny app containing full suite of disease associations: MaGE, Epigenetic Clocks and Clinical measurements

An R Shiny app aggregates the results of all Cox regression models, including survival, clinical and comparative analyses (HTTPS://HSR-RSC-PROD3.MAYO.EDU:1919/MAGE_DISEASE_ASSOC_SEP2026/). Users can visualize the findings of individual or multiple DNAm predictors for singular disease outcomes and groupings.

## Code availability

Fully annotated R and Python analysis scripts are available at https://github.com/dsimps1993/Methylation-associated-Gene-Expression-Predictors

## Acknowledgement

We thank all the Generation Scotland (GS) study participants and research team members who have contributed, and continue to contribute, to ongoing studies. This research was funded in part, by the Wellcome Trust grant 218492/Z/19/Z GS received core support from the Chief Scientist Office of the Scottish Government Health Directorates (CZD/16/6) and the Scottish Funding Council (HR03006). DNA methylation profiling of the GS samples was carried out by the Genetics Core Laboratory at the Wellcome Trust Clinical Research Facility, Edinburgh, UK, and was funded by the Medical Research Council UK, the Brain & Behavior Research Foundation (ref. 27404) and the Wellcome Trust (Wellcome Trust Strategic Award ‘STratifying Resilience and Depression Longitudinally’ ((STRADL) ref. 104036/Z/14/Z)). The authors also acknowledge the generous support of the Beverly foundation.

## COIs

R.E.M. is an advisor to the Epigenetic Clock Development Foundation and Optima Partners. S.C., N.K.L., and T.C. have intellectual property related to the research reported here, which has been reviewed by the Mayo Clinic Conflict of Interest Review Board and is being conducted in compliance with Mayo Clinic Conflict of Interest policies. A provisional patent has been filed for MaGE (US provisional 63/882,750).

## Author contributions

T.C., N.K.L., D.J.S., S.J.C., and C.M. all contributed to conceptualization of this work. All authors were involved in writing the manuscript. D.J.S., S.J.C., C.M., Y.S., T.A.W. and K.L. ran the computational analysis and data generation. R.E.M. and A.R. curated and provided access to Generation Scotland.

---

**Supplementary Figure 1.**
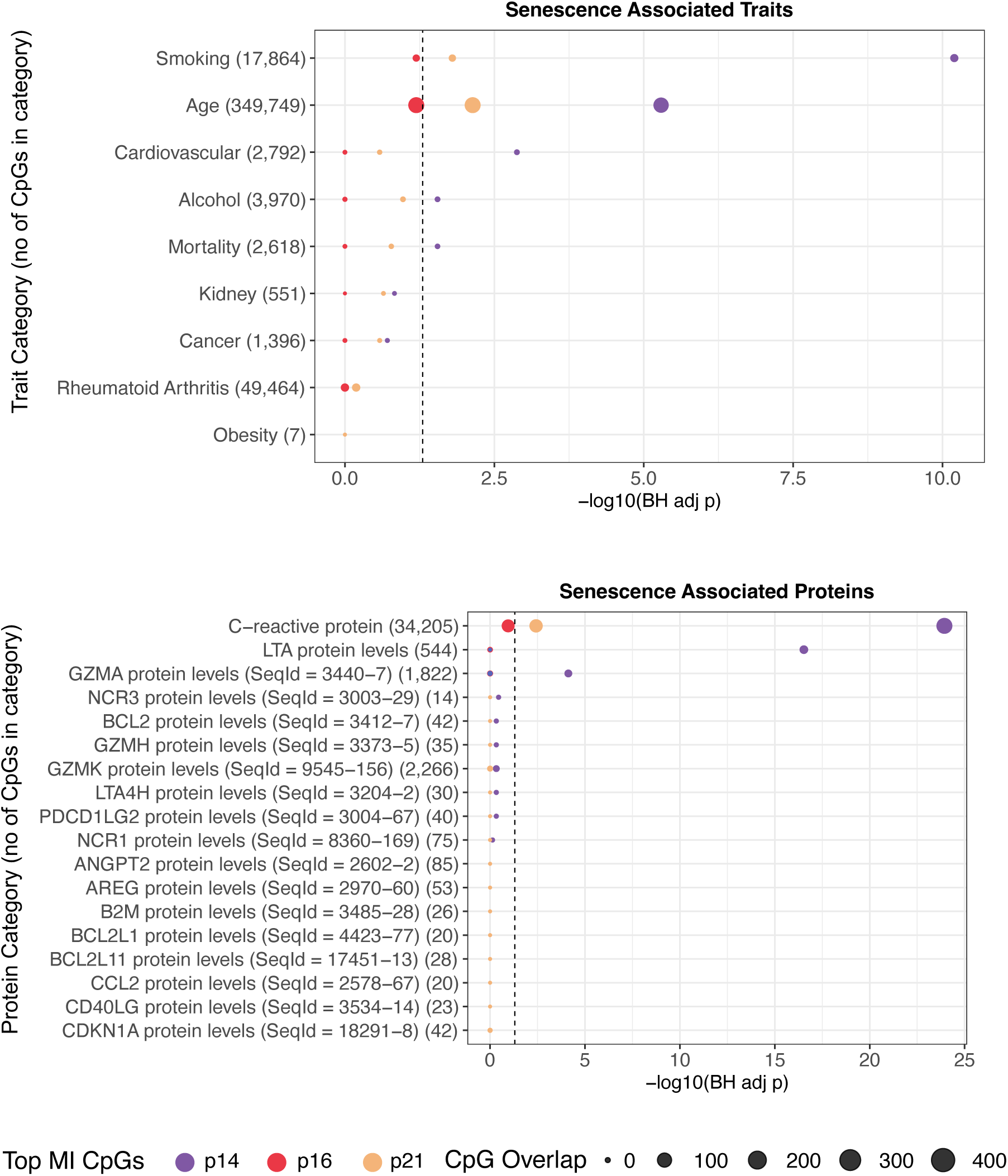
Enrichment of EWAS catalog terms with top CpGs associated with p14^ARF^, p16^INK4A^ or p21^CIP1^ expression. Phenotypic senescence traits (top) and senescence associated proteins (bottom). For the EWAS catalog enrichment analysis, the top 1000 CpGs with mutual information with each senescence-associated protein were filtered to CpGs present on both the EPIC and 450K array, resulting in 434, 515, and 503 CpGs for p14^ARF^, p16^INK4A^ and p21^CIP1^ respectively. These CpGs were tested for over-representation among CpGs associated with curated senescence-related trait categories (top) and a curated panel of 108 senescence related proteins (bottom) in blood-tissue EWAS studies using one-sided Fisher’s exact tests. The x-axis shows −log_10_(p-value) for each enrichment test, with the dashed vertical line indicating nominal significance (p = 0.05). Point color denotes the predictor, and point size reflects the number of overlapping CpGs between the predictor and the trait category. Diamond-shaped points indicate enrichments that remain significant after Benjamini-Hochberg correction (adjusted p < 0.05); circles indicate nominal or non-significant results. Parenthetical values on the y-axis denote the number of background CpGs associated with each trait category in the EWAS Catalogue.

**Supplementary Figure 2.**
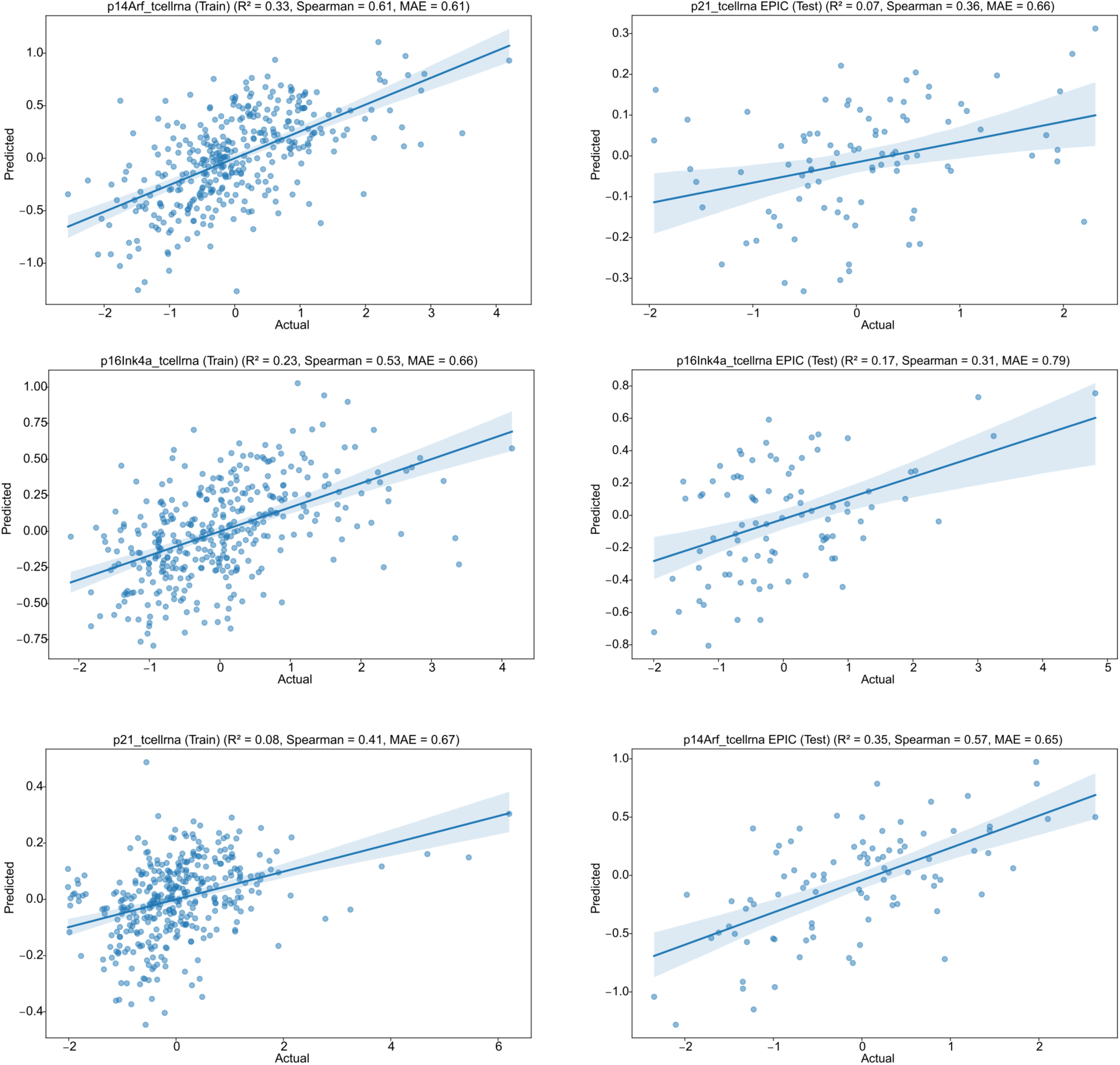
Senescence MaGE predictors (MaGE-p14; first row, MaGE-p16; second row, MaGE-p21; third row) applied to training (first column) and test datasets (second column). Actual expression (x-axis) is z-score normalized qPCR expression, predicted expression is z-score normalized MaGE output.

**Supplementary Figure 3.**
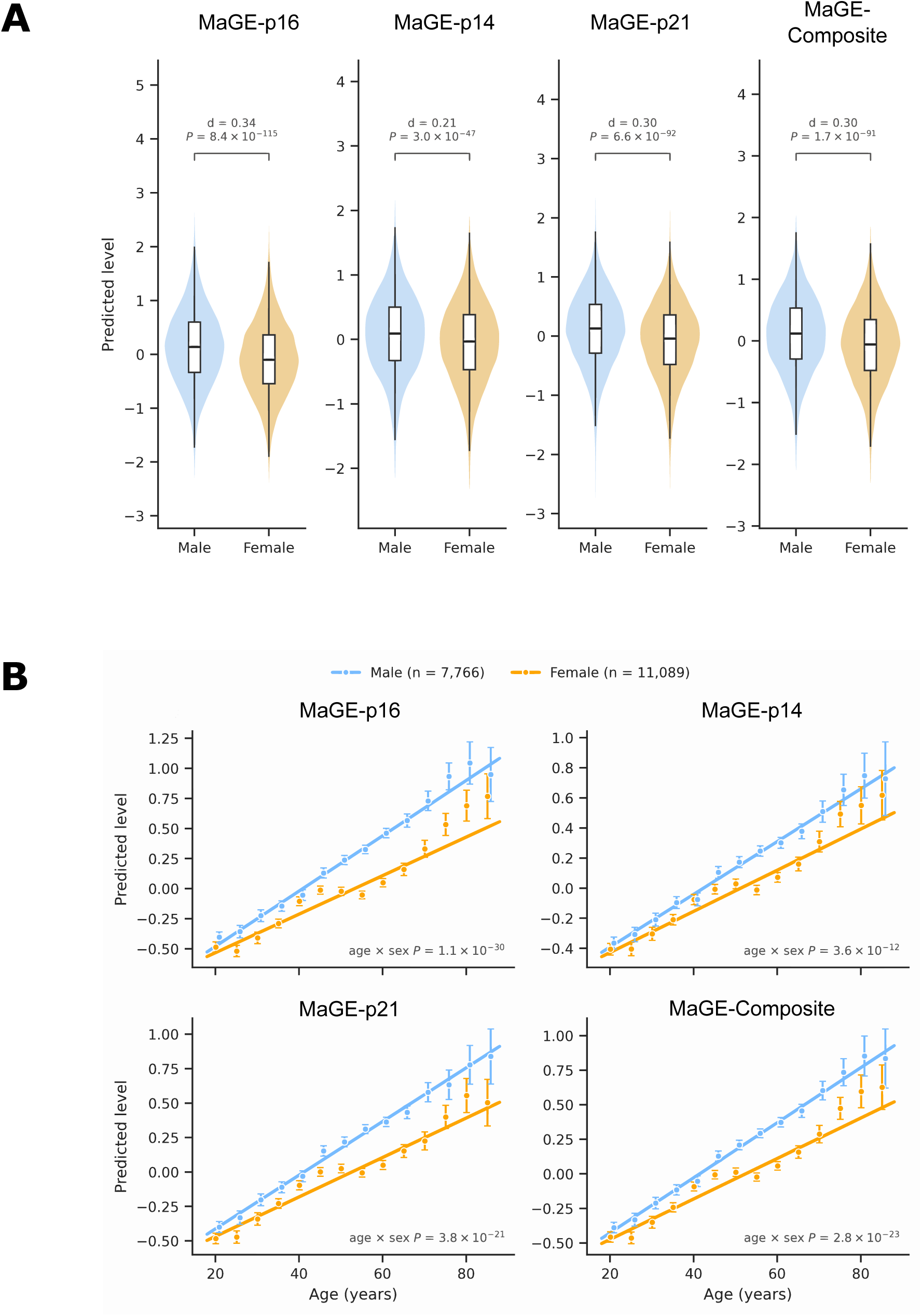
Predicted senescence MaGE is higher and rises faster with age in males than females. (**A**) Violin plot of predicted MaGE-p16, MaGE-p14, MaGE-p21 and MaGE-Composite output separated by gender (males; blue, n=7,766, females; orange, n = 11,089). Violins show the distribution of each score, scaled to equal width. Boxes show the median and interquartile range, and whiskers extend to 1.5× the interquartile range. P values are from a linear regression of each score on sex, with standard errors clustered by family to account for relatedness. d is Cohen’s d, the difference in means (male − female) divided by the pooled standard deviation. (**B**) Predicted MaGE-p16, MaGE-p14, MaGE-p21 and MaGE-Composite output against age for males (blue) and females (orange). Each marker is standardized to mean 0 and SD 1 across the analysis sample. Points are means in 5-year age bins, with error bars showing 95% confidence intervals. Lines are sex-specific fits from a linear model with a separate intercept and age slope for each sex. P values test whether the age slope differs between the sexes (the age × sex interaction). Standard errors are clustered by family to account for relatedness.

**Supplementary Figure 4.**
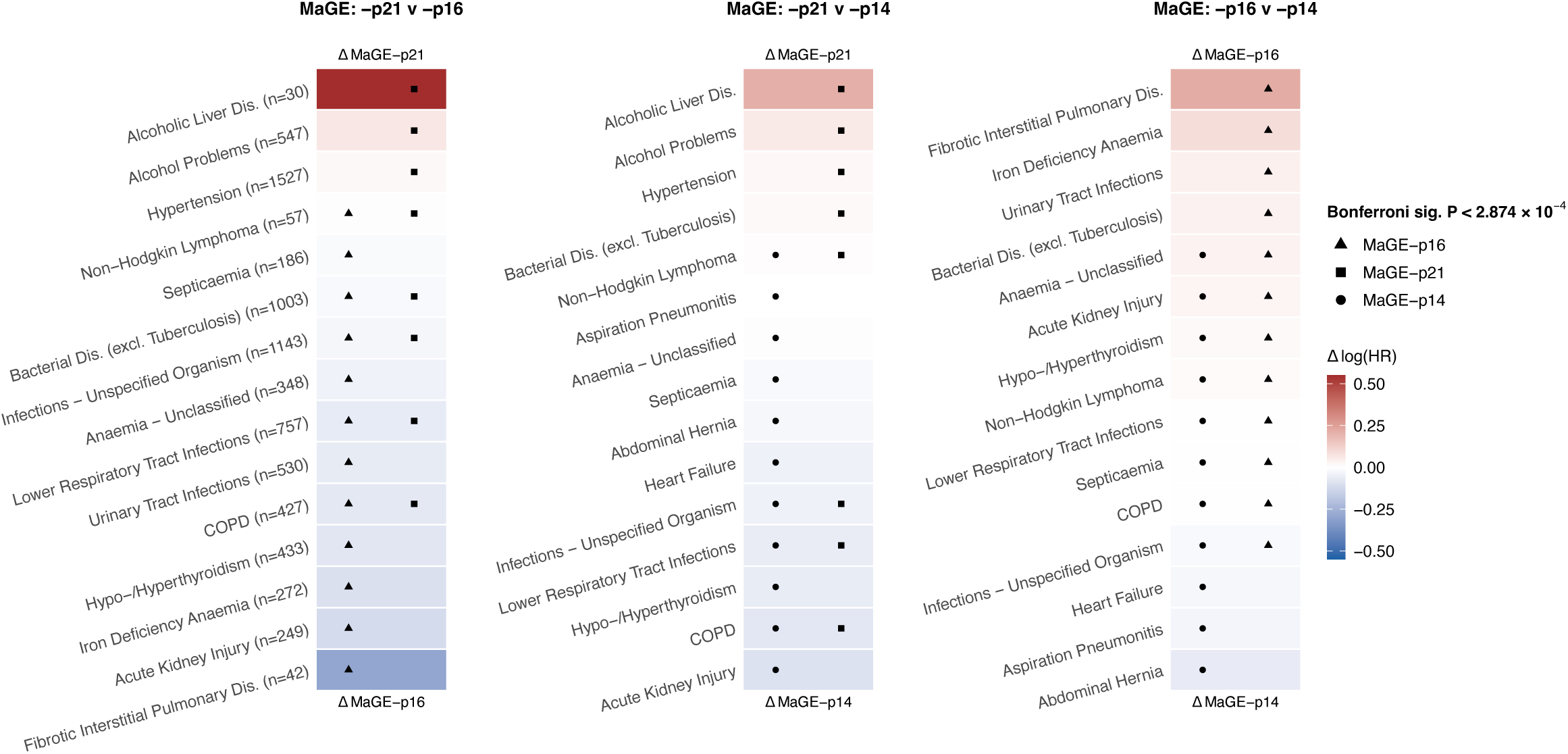
Differential disease associations between MaGE-p14, MaGE-p16, and MaGE-p21 predictors. Heatmaps of the pairwise difference in log(hazard ratio) [Δlog(HR)] between MaGE predictors for their associations with incident disease outcomes in Generation Scotland. Each panel compares two predictors as indicated by the axis labels (Δ MaGE-p21 vs Δ MaGE-p16, left; Δ MaGE-p21 vs Δ MaGE-p14, middle; Δ MaGE-p16 vs Δ MaGE-p14, right), with diseases ranked by the magnitude of delta. Red indicates a stronger association (higher log(HR)) for the predictor named first in each panel title, and blue indicates a stronger association for the predictor named second. Marker shapes denote which predictor(s) reached Bonferroni-corrected significance (P < 2.874 × 10⁻ ^4^) for that disease: triangle = MaGE-p16, square = MaGE-p21, circle = MaGE-p14. Case numbers (n) for each disease are shown in the left panel.

**Supplementary Figure 5.**
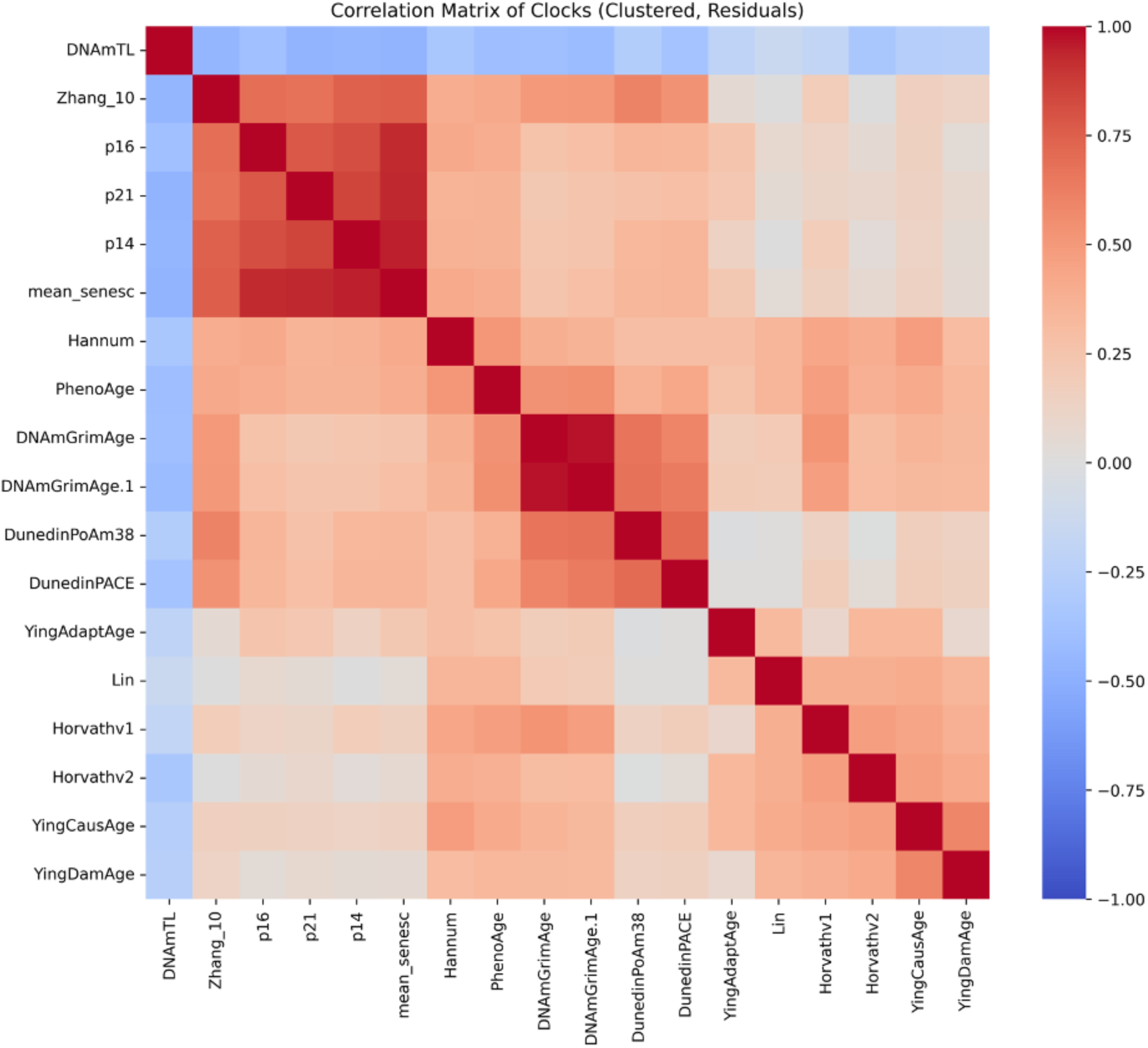
Cross correlation analysis of senescence MaGE predictor output and age acceleration (residuals) for 14 published epigenetic clocks, applied to Generation Scotland.

**Supplementary Figure 6.**
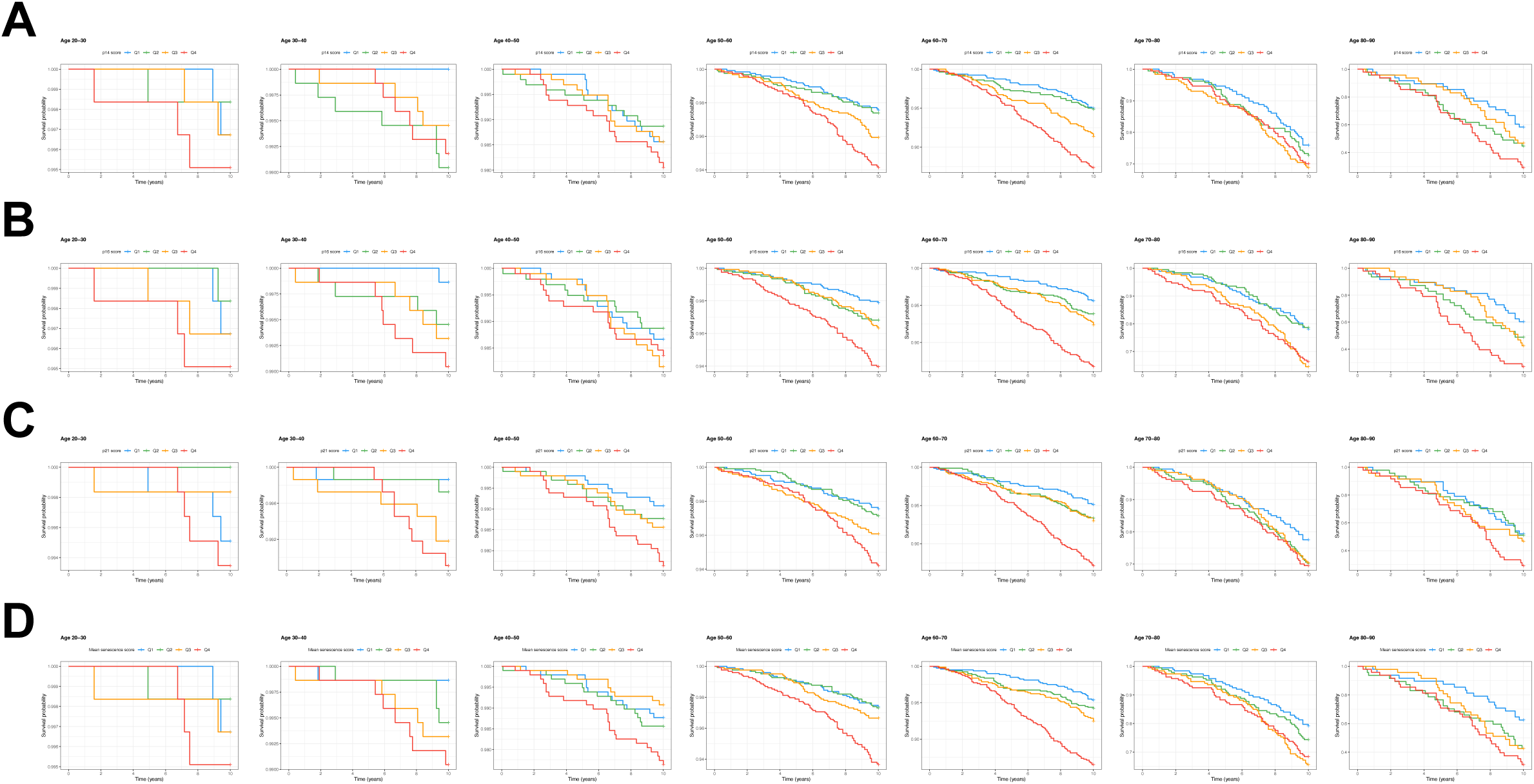
Kaplan-Meier curves of MaGE predictors applied to the Generation Scotland cohort, stratified by age groups. **(A)** MaGE-p14, **(B)** MaGE-p16, **(C)** MaGE-p21 and **(D)** MaGE-Composite.

**Supplementary Figure 7.**
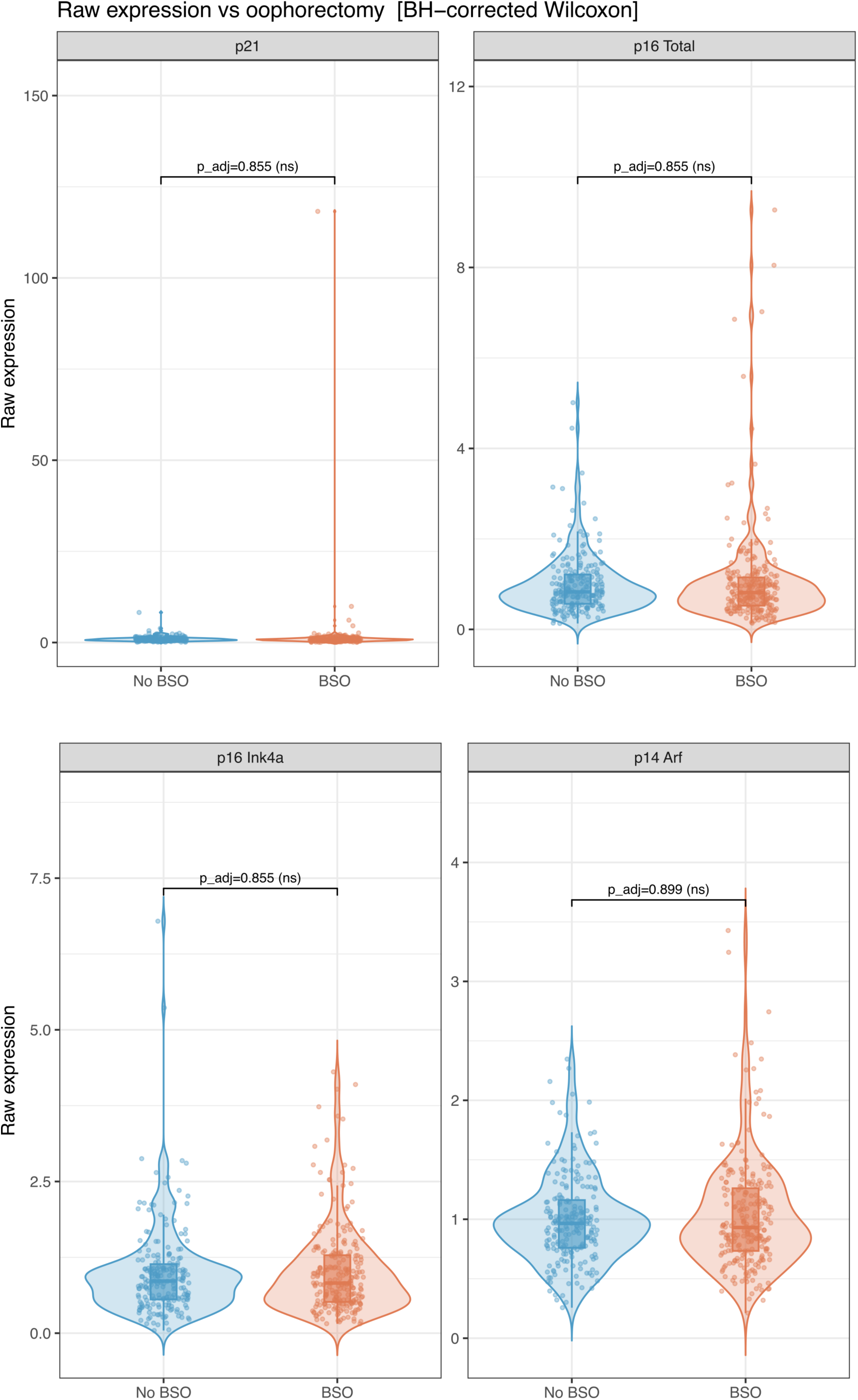
Raw qPCR expression values of p21, p16 total, p16^INK4A^ and p14^ARF^, stratified by bilateral salpingo-oophorectomy (BSO) status. Wilcoxon rank sum test was conducted between BSO and no-BSO (ns=not significant).

**Supplementary Table 1.** Summary statistics of Mayo Clinic Cohort Study of Oophorectomy and Aging-2 (MOA-2). Hormone therapy treatment was stratified by Bilateral Salpingo-Oophorectomy (BSO) status.

| **Variable** | **Overall**<br>N = 455<sup>1</sup> | **Test**<br>N = 92<sup>1</sup> | **Train**<br>N = 363<sup>1</sup> |
| --- | --- | --- | --- |
| **Oophorectomy Status** |  |  |  |
| BSO | 242 (53%) | 49 (53%) | 193 (53%) |
| No BSO | 213 (47%) | 43 (47%) | 170 (47%) |
| **Estrogen Therapy (by Age 50)** |  |  |  |
| No Estrogen | 251 (55%) | 53 (58%) | 198 (55%) |
| Estrogen Therapy | 204 (45%) | 39 (42%) | 165 (45%) |
| **Age at Visit 1** | 66.5 | 66.8 | 66.5 |
| **p14ARF Raw Expression** | 0.94 | 0.97 | 0.93 |
| **p16INK4A Raw Expression** | 0.85 | 0.83 | 0.86 |
| **p21CIP1 Raw Expression** | 0.84 | 0.88 | 0.83 |
| <sup>1</sup> n (%); Median |  |  |  |

